# Power calculations for detecting differences in efficacy of fecal microbiota donors

**DOI:** 10.1101/2020.04.16.20068361

**Authors:** Scott W. Olesen

**Affiliations:** OpenBiome, 2067 Massachusetts Avenue, Cambridge, MA 02140, USA

## Abstract

Fecal microbiota transplantation (FMT) is a recommended therapy for recurrent *Clostridioides difficile* infection and is being investigated as a potential therapy for dozens of other indications, notably inflammatory bowel disease. The immense variability in human stool, combined with anecdotal reports from FMT studies, have suggested the existence of “donor effects”, in which stool from some FMT donors is more efficacious than stool from other donors. In this study, simulated clinical trials were used to estimate the number of patients that would be required to detect donor effects under a variety of study designs. In most cases, reliable detection of donor effects required more than 100 patients treated with FMT. These results suggest that previous reports of donor effects need to be verified with results from large clinical trials and that patient biomarkers may be the most promising route to robustly identifying donor effects.

## Introduction

Fecal microbiota transplantation (FMT), the transfer of stool from healthy donors into ill patients, is a recommended therapy for multiply recurrent *Clostridioides difficile* infection, the most common hospital-acquired infection in the United States [1]. FMT is also being investigated as a potential therapy for dozens of other indications, including inflammatory bowel disease, metabolic diseases, cancer, and central nervous disorders [1, 2]. Although FMT’s molecular mechanisms for treating *C. difficile* infection are not fully understood, FMT’s efficacy for treating *C. difficile* has motivated its experimental use in these other areas [3].

Stool donors are selected by a process of exclusion designed to maximize patient safety. Candidate donors are excluded based on blood and stool tests for known pathogens, risk factors for pathogen carriage, and personal and family history of potentially microbiome-mediated diseases [4]. However, given the enormous complexity of stool –which includes bacteria, viruses, fungi, microbe-derived molecules, and host-derived molecules– and the variability of stool from person to person [5, 6], it stands to reason that different stool could have different ability to treat disease. Anecdotes from FMT research, particularly in ulcerative colitis [7, 8], have created interest in the possibility of “donor effects”, that is, in potential variability in the efficacy of different donors’ stool [9, 10, 11, 12].

A “donor effect” refers to the possibility that stool from some donors is more efficacious than stool from other donors for treating some indication. For example, some FMT studies have tested for the possibility that one donor produced more efficacious stool than the other donors in the study [13, 7, 8]. Others have tested for the possibility that donors with greater microbiota diversity, or a differential abundance of some microbial taxon, is more efficacious [14, 15, 16, 17, 18]. Finally, at least one study has tested for the possibility that the composition of a donor’s gut microbiota is associated with outcome of a patient treated with that donor’s stool [19]. If donor effects do exist —if some donors, or some particular stool is more efficacious than other stool— they would be crucial to improving FMT as a therapy and to clarifying FMT’s molecular mechanisms [10].

Although multiple studies have tested for donor effects, these tests were all performed post hoc. In no case was a search for a donor effect part of the experimental design. It remains unclear if we can expect today’s FMT studies, mostly 20 to 40 patients in size [2], to reliably determine if donor effects exist. A key barrier to discovering donor effects, then, is a lack of statistical power methodology. Here we expand on previous theoretical models of donor effects [9, 20] and use simulations to estimate FMT studies’ statistical power to detect donor effects.

## Methods

Four designs for detecting donor effects were investigated.

### Contingency table model

In this model, variations of which were used in two previous theoretical studies of donor effects [9, 20], donors are assumed to be efficacious (“good”) or inefficacious (“bad”) and the distribution of donor efficacies is bimodal. Donor effects are tested for using a contingency table of patients outcomes by their associated donor. This approach was designed to model the statistical tests used to search for donor effects in previous FMT studies [13, 8, 7].

Specifically, a fraction ϕ of donors, the “efficacious” donors, have an efficacy *ε*+ (i.e., each patient treated with stool from an efficacious donor has a probability *ε*+ of a positive outcome). The remaining proportion 1 − *ϕ* of donors, the “inefficacious” donors, have efficacy *ε*− < *ε*+. To simplify the model, we make the assumptions that 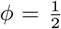 and the mean efficacy 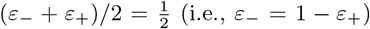, leaving only a single parameter Δ*ε* = *ε*_+_ *− ε*_*−*_ (Figure 1).

**Figure 1:**
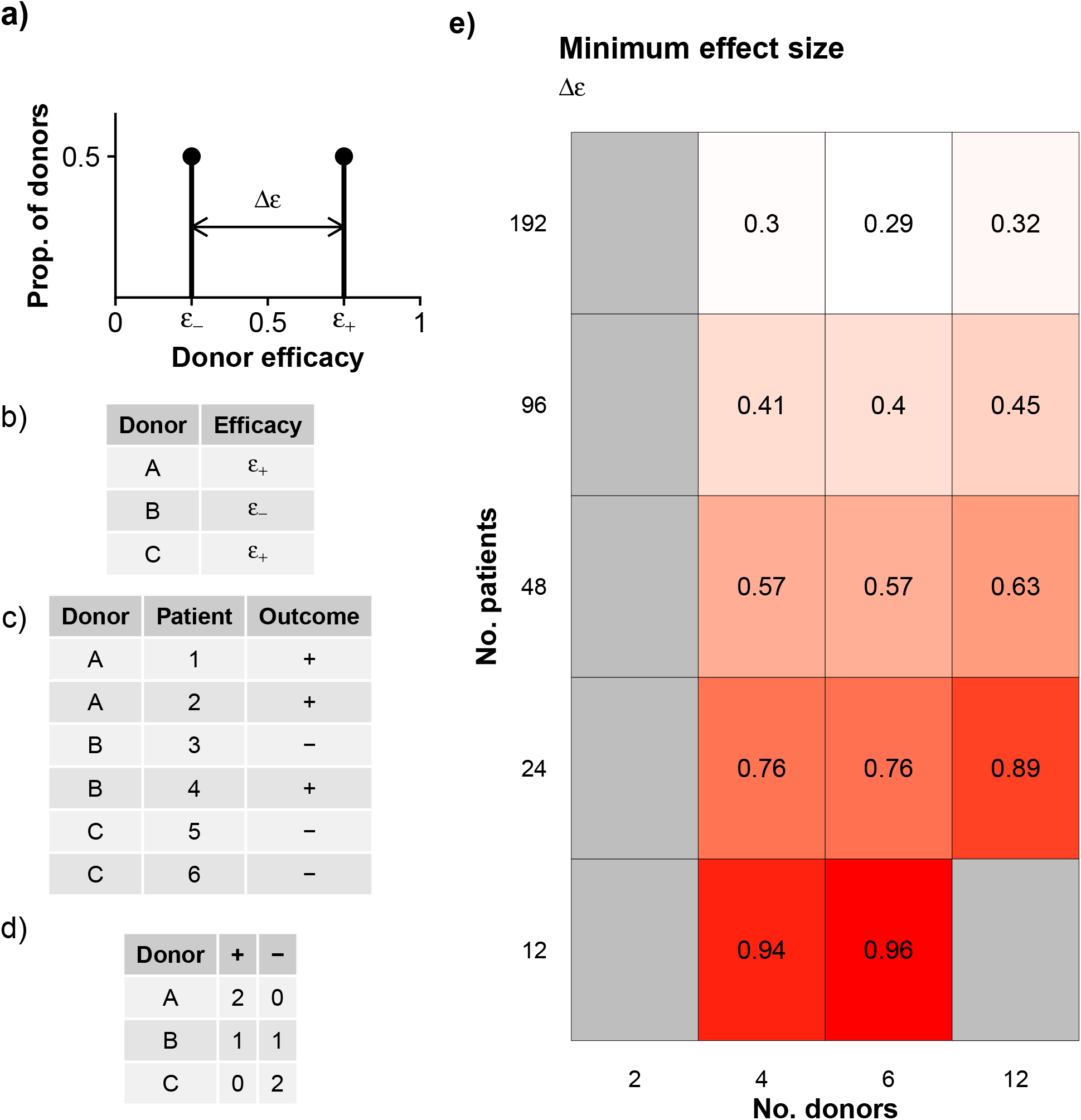
Contingency table model. a) In this model, half of donors have efficacy *ε*_+_ and the other half *ε*_*−*_, with mean efficacy 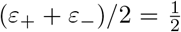. The effect size is Δ*ε* = *ε*_+_ *− ε*_*−*_. b) Donors’ efficacies are drawn from the distribution in a. c) Patient outcomes are drawn based on their associated donors’ efficacies. d) Statistical significance is assessed with a *χ*^2^ test on the contingency table of patient outcomes by donor. e) Minimum effect size required to reach 80% statistical power in simulated clinical trials.

Simulated clinical trials were run to determine the relationship between the effect size Δ*ε* and the statistical power of the study. In each simulated trial, *N*_*P*_ patients were evenly distributed among *N*_*D*_ donors. Each donor’s underlying efficacy, either *ε*_*−*_ or *ε*_+_, is randomly set according to ϕ. Each patient’s dichotomous outcome is randomly determined depending on the efficacy of their donor. Heterogeneity among the donors’ efficacies was tested for using a *χ*^2^ test with Yates’s correction on the *N*_*D*_ *×* 2 contingency table of patient outcomes by donor.

### Donor biomarker model

In this model, donor effects are detected by searching for an association between some continuous-valued donor biomarker and the associated patients’ outcomes. This approach was designed to model the statistical tests used to search for donor effects in previous FMT studies, where the biomarker in question is typically the donor’s gut microbiota community diversity or the abundance of particular microbial taxa in the donor [21].

Specifically, each donor has a continuous-valued biomarker *X*, drawn from a normal distribution with standard deviation *σ*, which determines that donors’ efficacy according to logit^*−*1^(*βX*). To simplify the model, we assume that donors with the mean biomarker values have efficacy 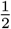. Then the model has one effect size *βσ*, which is the log odds ratio in donor efficacy per standard deviation increase in biomarker value. In other words, donor efficacies are log-normal distributed with shape parameter *βσ*. High *β* means that donors with different biomarkers have more distinct efficacies; high *σ* means that a sample of donors will have a wider range of biomarkers and efficacies.

For example, for *βσ* = 0, all donors have efficacy 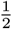. For *βσ* = 1, a donor with a biomarker one standard deviation above the mean has an efficacy of logit^*−*1^(1) *≈* 73%, while a donor one standard deviation below the mean has efficacy logit^*−*1^(*−*1) *≈* 27%. For *βσ ≫* 1, half the donors have 0% efficacy and the other half have 100% efficacy (i.e., the same distribution of efficacies as for the contingency table model for Δ*ε* = 1). Below the critical value 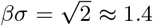, the distribution of donor efficacies is unimodal; above that value, it is bimodal.

In each simulated clinical trial, *N*_*P*_ patients receive FMT, each from a different donor, and their outcomes are simulated based on the randomly-sampled donor biomarkers *X* (Figure 2). A donor effect is detected using a Mann-Whitney test.

**Figure 2:**
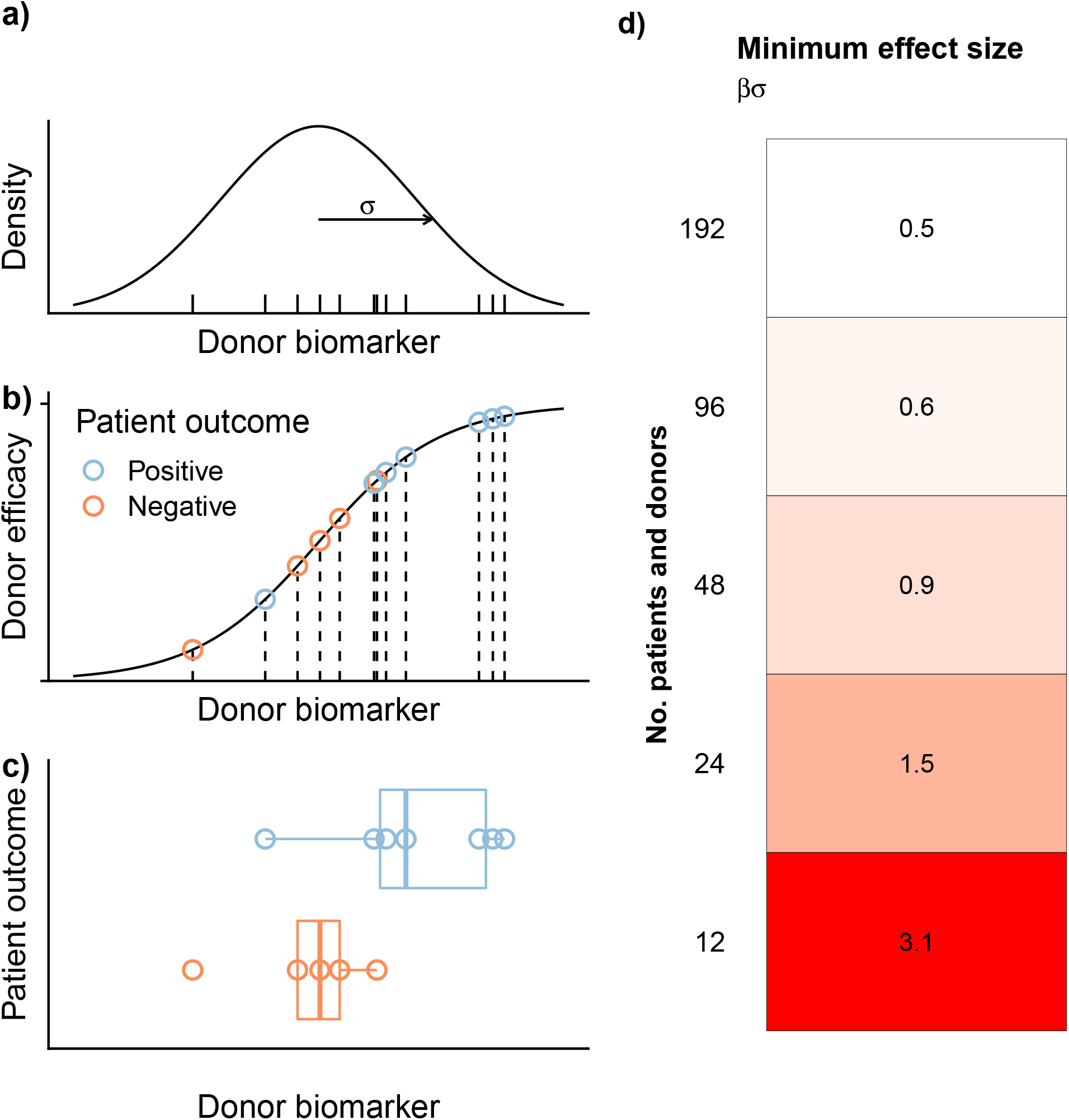
Donor biomarker model. a) Donor biomarkers (ticks) are drawn from a normal distribution (curve) with standard deviation *σ*. b) Donor efficacies (points) are determined based on their biomarkers *X* and the log odds parameter *β* according to *ε* = logit^*−*1^(*βX*). The effect size is *βσ*, the log odds ratio in donor efficacy per standard deviation of donor biomarker value. Patient outcomes (colors) are randomly determined based on their corresponding donors’ efficacies. c) A difference in donor biomarker values based on the outcomes of their corresponding patients is tested for with a Mann-Whitney test. c) Minimum effect size required to reach 80% statistical power in simulated clinical trials.

### Donor microbiota model

In this model, a donor effect is detected by looking for a separation in the donor gut microbiota composition by the donors’ associated patient outcomes. This approach was designed to model the investigation performed in Jacob *et al*. [19].

In this model, like in the contingency table model, donors have one of 2 efficacies, *ε*_*−*_ or *ε*_+_, and patients have dichotomous outcomes. Again, we make the simplifying assumptions that 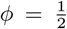 and 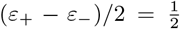. In this model, donor effects are detected based on separation of the microbiota of donors associated with patients who had positive outcomes, compared to the microbiota of donors associated with negative outcomes. A PERMANOVA test [22] checks for whether donors’ microbiota composition are associated with those donors’ associated patient outcomes.

This model has two relevant effect sizes. The first is the same effect size as in the contingency table model, Δ*ε*. The second is more subtle: how distinguishable are efficacious and inefficacious donors, in terms of their microbiota composition? To model “strong” versus “weak” separation in microbiota composition, data from previous case-control studies of diarrhea [23] and obesity [24] were used. The gut microbiota composition of cases with diarrhea are easily distinguishable from controls (in a random forest classifier, AUC *≈* 0.98), while obesity cases only weakly separate from healthy controls (AUC *≈* 0.69) [25].

In each simulation, a donor was assigned as efficacious or inefficacious, as in the contingency table model. Then, each efficacious donor was assigned a microbiota composition drawn at random from the controls in one of the two case-control studies. Each inefficacious donor was assigned a case’s microbiota composition (Figure 3). Next, 1 patient was assigned to each donor, and each patient’s outcome was determined at random according to the associated donor’s efficacy. Finally, a donor effect was searched for with a PERMANOVA test, comparing the donors’ microbiota condition on the patients’ outcomes.

**Figure 3:**
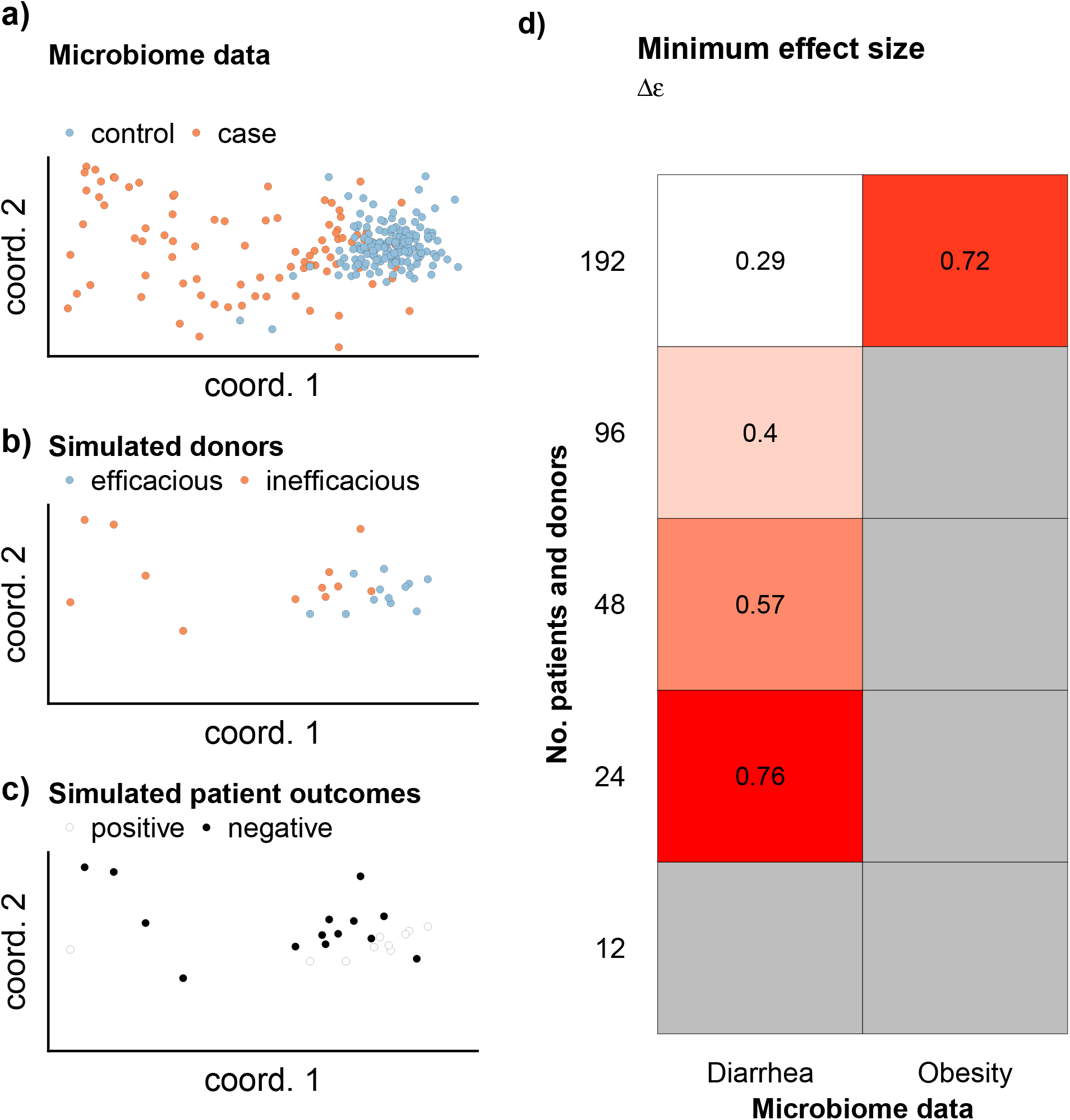
Donor microbiota model. Two donor efficacies, dichotomous patient outcomes, donor effect measured by separation of microbiota composition. a) An example ordination plot of the data from a microbiota case-control study. b) A subset of cases and controls, representing the simulated donors’ microbiota, are drawn, depending on the number of individuals in the simulated trial. c) Patient outcomes associated with each donor microbiota composition are drawn according to the efficacies *ε*_+_ and *ε*_*−*_. The effect size Δ*ε* determines the values *ε*_+_ and *ε*_*−*_ as in the contingency table model. A PERMANOVA test is run on the corresponding dissimilarity matrix of microbiota compositions to detect a donor effect. d) Minimum effect size required to reach 80% statistical power when using the contrast between cases and controls from microbiome studies as a proxy for the microbiome signature difference between efficacious and inefficacious controls.

Microbiota compositions were drawn from MicrobiomeHD [26], which processed 16S rRNA taxonomic marker gene sequences as described previously [25]. For the diarrhea study, non-*Clostridioides difficile* diarrhea patients were used as cases. For the obesity study, obese subjects were used as cases. The beta diversity matrix input into PERMANOVA was computed using the Bray-Curtis distance metric.

### Patient biomarker model

In this model, the donor effect is not restricted to efficacy of dichotomous donor outcomes. Instead, it is assumed that there is some continuous-valued biomarker measured in patients after the FMT, such as fecal calprotectin in the case of ulcerative colitis [27, 28] or a microbiome biomarker such as microbial engraftment. Heterogeneity in donors is detected using a Kruskal-Wallis test.

Specifically, each patient’s biomarker outcome is drawn from a normal distribution with standard deviation *σ*_*P*_, which is common to all patients, but centered on a mean value *µ*_*d*_ that is specific to the donor *d* used to treat that patient. The donor-associated values *µ*_*d*_ are themselves drawn from a hyperdistribution with standard deviation *σ*_*D*_ (Figure 4). The relevant effect size is the ratio *σ*_*D*_*/σ*_*P*_. For *σ*_*D*_*/σ*_*P*_ */gg*0, the variance in patients’ outcomes is due mostly to donor effects. For *σ*_*D*_*/σ*_*P*_ = 0, the variance is due solely to patient factors.

**Figure 4:**
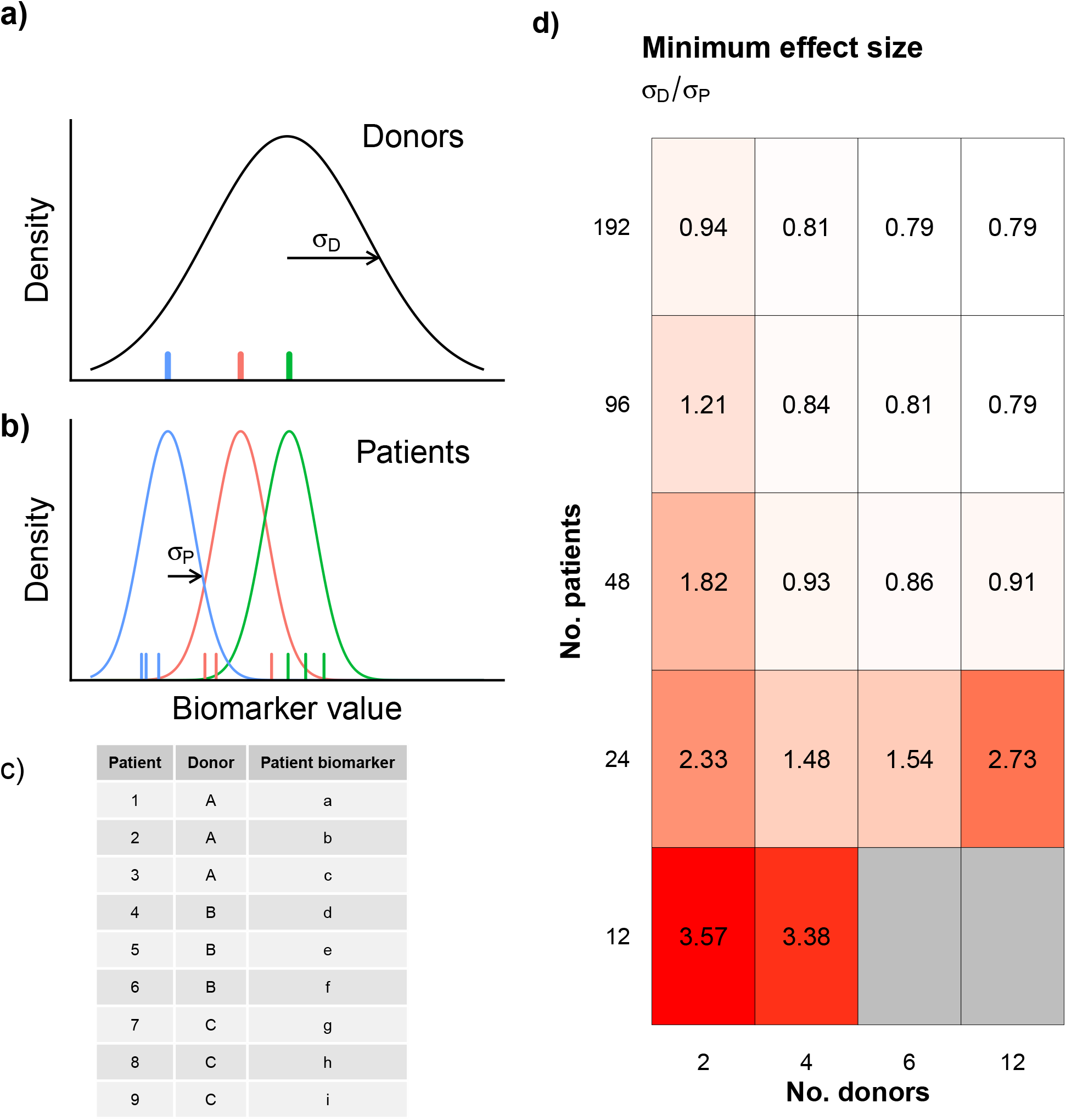
Patient biomarker model. Patient biomarker outcome. a) Each donor (colored ticks) has a mean patient biomarker value drawn from a hyperdistribution with standard deviation *σ*_*D*_ (black curve). b) Patients’ biomarkers (colored ticks) are drawn from distributions (colored curves), all with the same standard deviation *σ*_*P*_, centered on the mean of their associated donor. The effect size is *σ*_*D*_*/σ*_*P*_. c) Donor effects are detected by a Kruskal-Wallis test on patient biomarker values. d) Minimum effect size required to reach 80% statistical power in simulated clinical trials.

## Simulations

For all models, the number of patients was varied over 12, 24, 48, 96, and 196. For the contingency table and patient biomarker models, the number of donors was varied over 2, 4, 6, 8, and 12. For the donor biomarker and donor microbiota models, the number of donors was equal to the number of patients. For the contingency table and donor microbiota models, the effect size Δ*ε* was varied over [0, 1]. For the donor biomarker model, *βσ* was varied over [0, log 25]. For the patient biomarker model, *σ*_*D*_/(*σ*_*P*_ + *σ*_*D*_) was varied over [0, 1].

At 11 points along a grid of effect sizes in each model’s range, 1,000 simulations were conducted to compute the statistical power at the 0.05 confidence level. All calculations were performed using R (version 3.6.0) [29]. *χ*^2^ tests were formed using *chisq*.*test*, Mann-Whitney tests using *wilcox*.*test*, Kruskal-Wallis tests using *kruskal*.*test*, and PERMANOVA tests using the function *adonis* in the *vegan* package (version 2.5-6) [30]. Code is available at Zenodo (DOI: 10.5281/zenodo.3755048)

## Results

In the contingency table model, 80% statistical power was achieved in simulations similar to a typical FMT trial (i.e., with 24 patients), but the minimum effect size to achieve that power was Δ*ε*_+_ = 0.76 (Figure 1). In other words, the difference between efficacious and inefficacious donors had to be so great that on average 88% of patients assigned to efficacious donors had positive outcomes (i.e.,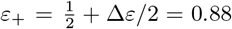), and only 12% of patients assigned to inefficacious donors had positive outcomes. As the number of patients in the simulated trials was increased, the minimum effect size to achieve 80% power decreased: for simulated trials with 192 patients receiving FMT, the minimum effect size was Δ*ε* = 0.3.

In the donor biomarker model, 80% statistical power was achieved with 24 patients and a minimum effect size of *βσ* = 1.5 (Figure 2). This effect size means that a donor with a biomarker one standard deviation above the mean has an efficacy of logit^*−*1^(1.5) *≈* 82%, and a donor one standard deviation below the mean has an efficacy of 18%. For 192 patients, this minimum effect size declined to 0.5 (i.e., donors one standard deviation above the mean have efficacy logit^*−*1^(0.5) *≈* 62%.

In the donor microbiota model, 80% statistical power was achieved with 24 patients when using the diarrhea case-control data as simulated efficacious and inefficacious microbiota, with a minimum effect size of Δ*ε* = 0.76 (i.e., efficacious donors have an efficacy 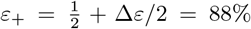; Figure 3). For 196 patients, the minimum effect size dropped to Δ*ε* = 0.29. When using the obesity case-control data, representing a more subtle difference between efficacious and inefficacious donors’ microbiota, 80% power was achieved only with 192 patients and Δ*ε* = 0.72.

In the patient biomarker model, 80% statistical power was achieved in simulations with 24 patients with a minimum effect size effect size *σ*_*D*_*/σ*_*P*_ = 3.4 (Figure 4). In other words, the variability in the mean patient biomarkers induced by different donors *σ*_*D*_ must be more than three times larger than the variability *σ*_*P*_ in patient biomarkers who receive FMT material from the same donor. For 192 patients, the minimum effect size declined to less than 0.8.

## Discussion

In this study, using 4 different models of donor effects, the minimum effect size required to achieve to achieve reasonable statistical power for a given number of patients were estimated. In the contingency table, donor biomarker, and donor microbiota models, 80% statistical power was achievable with 24 patients only when the effect sizes were implausibly large. These results suggest that current FMT trials, which typically include 20 to 40 patients treated with FMT, would seem unlikely to discover a true donor effect when using one of these approaches, and large clinical trials will be needed to verify previous reports of donors effects. The patient biomarker model, however, was more powerful than the other approaches.

### Contingency table model

In the clinical study [7] that motivated two previous modeling studies [20, 9], 38 patients were treated with FMT, the efficacy of the apparently efficacious donor was estimated as *ε*_+_ = 7/18 = 39%, and the efficacy of the remaining inefficacious donors was estimated as *ε*_*−*_ = 2/20 = 10%. In our simulations, a 30 percentage point difference in efficacy between the two classes of donors (i.e., Δ*ε* = 0.3) required more than 96 FMT-treated patients to be reliably detected.

### Donor biomarker model

In one clinical study [14], 13 patients were treated with FMT, and a significant difference in donor biomarkers (microbiota community diversity) between donors based on their associated patients’ outcomes was detected (Mann-Whitney test, *p* = 0.012). The results here suggest that, if such an effect were to be robustly detected in 12-patient trials, it would need to be implausibly large, with donors one standard deviation above the mean (i.e., 16% of donors) having an efficacy of at least 83%.

### Donor microbiota model

In one clinical study [19], 20 patients were treated with stool from 4 donors, with each patient receiving a mixture of 2 donors’ stool. A statisically significant separation in the mixtures’ microbiota composition based on the associated patients’ outcomes was detected (PERMANOVA, *p* = 0.044). By contrast, the results from this study suggest that reliable detection of such an effect in 24-patient studies would require both that efficacious and inefficacious donors have markedly different microbiota compositions (as different as diarrhea patients from healthy controls) and that their efficacies were implausibly distinct (Δ*ε* = 0.76, or 88% vs. 12% efficacy). These results broadly accord with previously work showing that microbiome studies are unlikely to robustly detect individual taxa mediating FMT patients’ outcomes [31].

### Patient biomarker model

The term “donor effects” has mostly been restricted to referring to differences in donor efficacy. However, finding any robust difference among donors in the effect that FMT has on patients would be helpful for understanding the molecular mechanism of FMT [10]. In the case of fecal calprotectin as a patient biomarker in inflammatory bowel disease, it appears that the variability in the biomarker among patients with the same disease severity is comparable to the variability in the mean biomarkers for different severities [28, 27]. In other words, if FMT from some donors could reliably move patients from severe to mild disease (i.e., *σ*_*D*_ *≈ σ*_*P*_), then the results of these simulations suggest that a donor effect could feasibly be detected with trials with as few as 48 patients. Furthermore, donor effects could feasibly be detected in animal studies when there is a relevant biomarker, even if the animal does not reach a clinical endpoint in the sense of a human clinical trial [32].

### Strengths and limitations

The key strength of this study was that it used straightforward mathematical models to make generous estimates of the statistical power of study designs that could detect donor effects. The key limitation of this study is that it does not account for the many sources of variance that arise in a clinical trial, most notably patient diagnoses and comorbidities. Thus, the estimates in this study should be taken as proof-of-concept only. In fact, additional variance coming from variability between patients, or from attempting an analysis using data from multiple clinical trials, will only increase the number of patients required to detect these effects.

## Conclusions

Given the large number of patients that would be required to prospectively detect a donor effect, *post hoc* detections of donor effects in small clinical trials should be verified with large clinical trials. The most promising path toward identifying donor effects, and using those effects to improve microbial therapeutics, appears to lie in using patient biomarkers, rather than donor biomarkers or in patient outcomes alone.

Clinical trialists should take care, however, that different approaches to testing for donor effects require mutually contradictory designs. For example, if a test will be run on patient biomarkers, then 2 to 6 donors should be used to maximize the study’s power to detect a donor effect. Using a different donor for every patient completely precludes the use of an Kruskal-Wallis or ANOVA test. However, if a donor effect will be detected via an association between donor biomarkers and patient outcomes, then a different donor should be used with each patient to maximize the study’s power. In that case, using the same donor to treat more than one patient will only make the subsequent analysis more complex and less powerful.

## Data Availability

Code to reproduce the results is available at Zenodo (DOI: 10.5281/zenodo.3755048).

## Acknowledgements

M. Santiago, Y. Gerardin, and E. Langner for helpful comments.

## References

[1] Jessica R Allegretti, Benjamin H Mullish, Colleen Kelly, and Monika Fischer. The evolution of the use of faecal microbiota transplantation and emerging therapeutic indications. The Lancet, 394(10196):420–431, August 2019.

[2] Scott W Olesen, Pratik Panchal, Justin Chen, Shrish Budree, and Majdi Osman. Global disparities in faecal microbiota transplantation research. The Lancet Gastroenterology & Hepatology, 5(3):241, March 2020.

[3] Alexander Khoruts and Michael J. Sadowsky. Understanding the mechanisms of faecal microbiota transplantation. Nature Reviews Gastroenterology & Hepatology, 13(9):508–516, June 2016.

[4] Giovanni Cammarota, Gianluca Ianiro, Colleen R Kelly, Benjamin H Mullish, Jessica R Allegretti, Zain Kassam, Lorenza Putignani, Monika Fischer, Josbert J Keller, Samuel Paul Costello, and et al. International consensus conference on stool banking for faecal microbiota transplantation in clinical practice. Gut, 68(12):2111–2121, Sep 2019.

[5] Lawrence A. David, Corinne F. Maurice, Rachel N. Carmody, David B. Gootenberg, Julie E. Button, Benjamin E. Wolfe, Alisha V. Ling, A. Sloan Devlin, Yug Varma, Michael A. Fischbach, and et al. Diet rapidly and reproducibly alters the human gut microbiome. Nature, 505(7484):559–563, Dec 236 2013.

[6] Peter J. Turnbaugh, Ruth E. Ley, Micah Hamady, Claire M. Fraser-Liggett, Rob Knight, and Jeffrey I. Gordon. The human microbiome project. Nature, 449(7164):804–810, Oct 2007.

[7] Paul Moayyedi, Michael G. Surette, Peter T. Kim, Josie Libertucci, Melanie Wolfe, Catherine Onischi, David Armstrong, John K. Marshall, Zain Kassam, Walter Reinisch, and et al. Fecal microbiota transplantation induces remission in patients with active ulcerative colitis in a randomized controlled trial. Gastroenterology, 149(1):102–109.e6, Jul 2015.

[8] Sudarshan Paramsothy, Michael A Kamm, Nadeem O Kaakoush, Alissa J Walsh, Johan van den Bogaerde, Douglas Samuel, Rupert W L Leong, Susan Connor, Watson Ng, Ramesh Paramsothy, and et al. Multidonor intensive faecal microbiota transplantation for active ulcerative colitis: a randomised placebo-controlled trial. The Lancet, 389(10075):1218–1228, Mar 2017.

[9] Scott W Olesen, Thomas Gurry, and Eric J Alm. Designing fecal microbiota transplant trials that account for differences in donor stool efficacy. Statistical Methods in Medical Research, 27(10):2906–2917, Feb 2017.

[10] Scott W. Olesen, McKenzie M. Leier, Eric J. Alm, and Stacy A. Kahn. Searching for superstool: maximizing the therapeutic potential of fmt. Nature Reviews Gastroenterology & Hepatology, 15(7):387–388, Apr 2018.

[11] Brooke C. Wilson, Tommi Vatanen, Wayne S. Cutfield, and Justin M. O’Sullivan. The super-donor phenomenon in fecal microbiota transplantation. Frontiers in Cellular and Infection Microbiology, 9, Jan 2019.

[12] Lito E. Papanicolas, David L. Gordon, Steve L. Wesselingh, and Geraint B. Rogers. Improving risk–benefit in faecal transplantation through microbiome screening. Trends in Microbiology, January 2020.

[13] Noortje G. Rossen, Susana Fuentes, Mirjam J. van der Spek, Jan G. Tijssen, Jorn H.A. Hartman, Ann Duflou, Mark Löwenberg, Gijs R. van den Brink, Elisabeth M.H. Mathus-Vliegen, Willem M. de Vos, and et al. Findings from a randomized controlled trial of fecal transplantation for patients with ulcerative colitis. Gastroenterology, 149(1):110–118.e4, Jul 2015.

[14] Severine Vermeire, Marie Joossens, Kristin Verbeke, Jun Wang, Kathleen Machiels, João Sabino, Marc Ferrante, Gert Van Assche, Paul Rutgeerts, and Jeroen Raes. Donor species richness determines faecal microbiota transplantation success in inflammatory bowel disease. Journal of Crohns and Colitis, 10(4):387–394, October 2015.

[15] P. Kump, P. Wurm, H. P. Gröchenig, H. Wenzl, W. Petritsch, B. Halwachs, M. Wagner, V. Stadlbauer, A. Eherer, K. M. Hoffmann, A. Deutschmann, G. Reicht, L. Reiter, P. Slawitsch, G. Gorkiewicz, and C. Hogenauer. The taxonomic composition of the donor intestinal microbiota is a major factor influencing the efficacy of faecal microbiota transplantation in therapy refractory ulcerative colitis. Alimentary Pharmacology & Therapeutics, 47(1):67–77, October 2017.

[16] Alka Goyal, Andrew Yeh, Brian R Bush, Brian A Firek, Leah M Siebold, Matthew Brian Rogers, Adam D Kufen, and Michael J Morowitz. Safety, clinical response, and microbiome findings following fecal microbiota transplant in children with inflammatory bowel disease. Inflammatory Bowel Diseases, 24(2):410–421, January 2018.

[17] Shinta Mizuno, Tatsuhiro Masaoka, Makoto Naganuma, Taishiro Kishimoto, Momoko Kitazawa, Shunya Kurokawa, Moeko Nakashima, Kozue Takeshita, Wataru Suda, Masaru Mimura, Masahira Hattori, and Takanori Kanai. Bifidobacterium-rich fecal donor may be a positive predictor for successful fecal microbiota transplantation in patients with irritable bowel syndrome. Digestion, 96(1):29–38, 2017.

[18] Atsushi Nishida, Hirotsugu Imaeda, Masashi Ohno, Osamu Inatomi, Shigeki Bamba, Mitsushige Sugimoto, and Akira Andoh. Efficacy and safety of single fecal microbiota transplantation for japanese patients with mild to moderately active ulcerative colitis. Journal of Gastroenterology, 52(4):476–482, October 2016.

[19] Vinita Jacob, Carl Crawford, Shirley Cohen-Mekelburg, Monica Viladomiu, Gregory G. Putzel, Yecheskel Schneider, Fatiha Chabouni, Sarah O’Neil, Brian Bosworth, Viola Woo, and et al. Single delivery of high-diversity fecal microbiota preparation by colonoscopy is safe and effective in increasing microbial diversity in active ulcerative colitis. Inflammatory Bowel Diseases, 23(6):903–911, Jun 2017.

[20] Abbas Kazerouni and Lawrence M. Wein. Exploring the efficacy of pooled stools in fecal microbiota transplantation for microbiota-associated chronic diseases. PLOS ONE, 12(1):e0163956, Jan 2017.

[21] Karen S.W. Leong, Justin M. O’Sullivan, José G.B. Derraik, and Wayne S. Cutfield. Gut microbiome transfer – finding the perfect fit. Clinical Endocrinology, March 2020.

[22] Marti J. Anderson. A new method for non-parametric multivariate analysis of variance. Austral Ecology, 26(1):32–46, Feb 2001.

[23] Alyxandria M. Schubert, Mary A. M. Rogers, Cathrin Ring, Jill Mogle, Joseph P. Petrosino, Vincent B. Young, David M. Aronoff, and Patrick D. Schloss. Microbiome data distinguish patients with clostridium difficile infection and non-c. difficile-associated diarrhea from healthy controls. mBio, 5(3), May 2014.

[24] Julia K. Goodrich, Jillian L. Waters, Angela C. Poole, Jessica L. Sutter, Omry Koren, Ran Blekhman, Michelle Beaumont, William Van Treuren, Rob Knight, Jordana T. Bell, and et al. Human genetics shape the gut microbiome. Cell, 159(4):789–799, Nov 2014.

[25] Claire Duvallet, Sean M. Gibbons, Thomas Gurry, Rafael A. Irizarry, and Eric J. Alm. Meta-analysis of gut microbiome studies identifies disease-specific and shared responses. Nature Communications, 8(1), Dec 2017.

[26] Claire Duvallet, Sean Gibbons, Thomas Gurry, Rafael Irizarry, and Eric Alm. Microbiomehd: the human gut microbiome in health and disease [data set]. zenodo.

[27] Ashish Kumar Jha, Madhur Chaudhary, Vishwa Mohan Dayal, Amarendra Kumar, Sanjeev Kumar Jha, Praveen Jha, Shubham Purkayastha, and Ravish Ranjan. Optimal cut-off value of fecal calprotectin for the evaluation of ulcerative colitis: An unsolved issue? JGH Open, 2(5):207–213, Aug 2018.

[28] Shuhei Fukunaga, Kotaro Kuwaki, Keiichi Mitsuyama, Hidetoshi Takedatsu, Shinichiro Yoshioka, Hiroshi Yamasaki, Ryosuke Yamauchi, Atsushi Mori, Tatsuyuki Kakuma, Osamu Tsuruta, and et al. Detection of calprotectin in inflammatory bowel disease: Fecal and serum levels and immunohistochemical localization. International Journal of Molecular Medicine, Nov 2017.

[29] R Core Team. R: A Language and Environment for Statistical Computing. R Foundation for Statistical Computing, Vienna, Austria, 2019.

[30] Jari Oksanen, F. Guillaume Blanchet, Michael Friendly, Roeland Kindt, Pierre Legendre, Dan McGlinn, Peter R. Minchin, R. B. O’Hara, Gavin L. Simpson, Peter Solymos, M. Henry H. Stevens, Eduard Szoecs, and Helene Wagner. vegan: Community Ecology Package, 2019. R package version 2.5-6.

[31] Claire Duvallet, Caroline Zellmer, Pratik Panchal, Shrish Budree, Majdi Osman, and Eric J. Alm. Framework for rational donor selection in fecal microbiota transplant clinical trials. PLOS ONE, 14(10):e0222881, October 2019.

[32] Takeshi Tanoue, Satoru Morita, Damian R. Plichta, Ashwin N. Skelly, Wataru Suda, Yuki Sugiura, Seiko Narushima, Hera Vlamakis, Iori Motoo, Kayoko Sugita, Atsushi Shiota, Kozue Takeshita, Keiko Yasuma-Mitobe, Dieter Riethmacher, Tsuneyasu Kaisho, Jason M. Norman, Daniel Mucida, Makoto Suematsu, Tomonori Yaguchi, Vanni Bucci, Takashi Inoue, Yutaka Kawakami, Bernat Olle, Bruce Roberts, Masahira Hattori, Ramnik J. Xavier, Koji Atarashi, and Kenya Honda. A defined commensal consortium elicits CD8 T cells and anti-cancer immunity. Nature, 565(7741):600–605, January 2019.

